# Higher Perceived Stress during the COVID-19 pandemic increased Menstrual Dysregulation and Menopause Symptoms

**DOI:** 10.1101/2022.07.30.22278213

**Authors:** Romina Garcia de leon, Alexandra Baaske, Arianne Y Albert, Amy Booth, C. Sarai Racey, Shanlea Gordon, Laurie W. Smith, Anna Gottschlich, Manish Sadarangani, Angela Kaida, Gina S. Ogilvie, Lori A. Brotto, Liisa A.M. Galea

**Author notes:** Correspondence can be addressed to: Dr. Liisa Galea, Djavad Mowafaghian Centre for Brain Health, University of British Columbia, 2215 Wesbrook Mall, Vancouver, BC, Canada, V6T 1Z3. **Statements and Declarations**. <u>Ethical Approval</u> Ethical approval was received from the BC Children’s and Women’s Research Ethics board (H20–01421). All methods performed as a part of this study were in accordance with the UBC Research Ethics Board guidelines. <u>Informed consent</u> to participate was obtained from participants. <u>Data Availability</u> Data cannot be shared publicly because of ethical restrictions. Data are available from the UBC Research Ethics Board (contact via) for researchers who meet the criteria for access to confidential data. <u>Author Contributions:</u> All authors contributed to the study conception and design. Material preparation, data collection and analysis were performed by Arianne Albert, Amy Booth, Shanlea Gordon, Alexandra Baaske and Romina Garcia de Leon. The first draft of the manuscript was written by Romina Garcia de Leon, Alexandra Baaske, and Liisa Galea and all authors commented on previous versions of the manuscript. All authors read and approved the final manuscript.

## Abstract

**Objective:** The increased stress the globe has experienced with the COVID-19 pandemic has affected mental health, disproportionately affecting women. However, how perceived stress in the first year affected menstrual and menopausal symptoms has not yet been investigated.

**Methods:** Residents in British Columbia, Canada, were surveyed online as part of the COVID-19 Rapid Evidence Study of a Provincial Population-Based Cohort for Gender and Sex (RESPPONSE). A subgroup (n=4171) who were assigned female sex at birth (age 25-69) and were surveyed within the first 6-12 months of the pandemic (August 2020-February 2021), prior to the widespread rollout of vaccines, were retrospectively asked if they noticed changes in their menstrual or menopausal symptoms, as well as completing validated measures of stress, depression, and anxiety.

**Results:** We found that 27.8% reported menstrual cycle disturbances and 6.7% reported increased menopause symptoms. Those who scored higher on perceived stress, depression, and anxiety scales were more likely to have reproductive cycle disturbances. Free text responses revealed that reasons for disturbances were perceived to be related to the pandemic.

**Conclusions:** The COVID-19 pandemic has highlighted the need to research women’s health issues, such as menstruation. Our data indicates that in the first year of the pandemic, almost a third of the menstruating population reported disturbances in their cycle, which is approximately two times higher than in non-pandemic situations and four times higher than any reported changes in menopausal symptoms across that first year of the pandemic.

**Summary Sentences:** Women+ with higher anxiety, depression or perceived stress scores during the first year of the pandemic were more likely to have experienced menstrual cycle phase disturbance or menopausal status disruption. Younger women were particularly prone to disturbances in their reproductive cycles.

---

The COVID-19 pandemic led to closures of public places, strict health regulations, and limited social interactions. These changes, along with the uncertainty of the pandemic, resulted in increased perceived levels of stress, depression, and anxiety^1,2^. Women and gender diverse individuals exhibited greater indices of distress, including heightened risk for mental health disorders than men throughout the COVID-19 pandemic ^1,3,4^. Despite these findings, there has been minimal attention paid to the effects of sex and gender in COVID-19 studies^5–7^. There has been a lack of research on the specific impact of SARS-CoV-2 vaccines, and pandemic response on female health^8^ which has contributed to alarming anecdotal reports of menstrual cycle irregularities following COVID-19 vaccination on social media^9^ These reported incidents underscored the importance of studying female cycles, which are now being closely examined in relation to COVID-19 vaccination^7,10,11^. However, there remains a need to examine the overall impact of the pandemic response on female menstrual cycles. Menstrual cycle irregularities, perimenopause, and menopausal disturbances can be important indicators of overall physical health^12^. Menstrual cycle irregularity is linked to metabolic dysfunction^13^ and increased risk of cardiovascular disease^14^. Furthermore, vasomotor menopausal symptoms are related to white matter hyperintensities and cognitive disturbances^15^. Exposure to stress affects the hypothalamic pituitary gonadal (HPG) axes^16^ as higher perceived stress is correlated with menopausal symptoms^17,18^ and menstrual dysregulation^19^. Thus, it is not surprising that the COVID-19 pandemic and the associated increase in stress has led to reports of disturbances to menstrual cycles^20^. Premenopausal people have reported irregularities in their menstrual cycles across different phases of the COVID-19 pandemic^21^,consistent with the literature indicating that heightened perceived stress is a predictor for menstrual irregularities^22^. Prolonged stress is also associated with an increased risk for psychiatric disorders^23^ and females reported higher depressive symptoms than males during the COVID-19 pandemic^1,24^. Moreover, studies show that depressed mood is associated with menstrual irregularities^25^, meaning that pandemic-related stress may be affecting mental health and menstrual irregularities simultaneously or sequentially. Given that stress and menstrual irregularities are connected^22^, and may play a role in mental health disturbances^25^, it is important to study these relationships. Heightened stress would likely have an impact on menstrual/menopausal irregularities, but to our knowledge no study to date has examined these irregularities in relation to psychosocial outcomes such as perceived stress, anxiety, or depression scores, and other factors that may play a role in perceived stress (age, number of children) within the context of the COVID-19 pandemic. We hypothesized that females would report an increase in the number of menstrual and menopausal symptoms during the first year of the pandemic, and secondly that these disturbances would be associated with stress, anxiety and depression levels.

## Methods

### Participant Recruitment

The Rapid Evidence Study of a Provincial Population Based COhort for GeNder and SEx (RESPPONSE) was led by the Women’s Health Research Institute in British Columbia (BC). All participants provided informed consent prior to participation in RESPPONSE. Ethics approval was received from The BC Children’s and Women’s Research Ethics Board (H20-01421). Between mid-August 2020 and March 1, 2021, participants were recruited^26^ Og. Survey responses were collected anonymously, with the exception of postal code. All respondents who completed the survey were invited to enter a draw to win a $100 e-gift card. The survey was open to residents of BC aged 25 to 69 years of all sexes and genders. However, only participants who were assigned female sex at birth were eligible for this particular analysis (n=5608). For analyses pertaining to menstrual changes, eligibility was restricted to: female sex, not post-menopausal, not pregnant, first six months postpartum, and not on hormonal suppression drugs (n=1866). In addition, we also conducted sensitivity analyses for menstrual changes to those people under 40 to avoid conflation with possible symptoms of perimenopause (n=810). For analyses of menopausal symptoms, eligibility was restricted to female sex and those who responded that they were postmenopausal (n=2305). In addition, we also conducted sensitivity analyses for menopause status to those people over 50 (n=1978) to avoid conflation with possible symptoms of perimenopause.

### Survey Design and Measures

The survey was tested for face validity, pilot tested, and implemented using REDCap (Research Electronic Data Capture)^27^. The survey consisted of multiple modules, with the present analysis focused on female reproductive health questions surrounding menstrual cycle and menopause symptoms. All survey participants that were premenopausal were asked ”Since the COVID-19 pandemic and subsequent public health measures in mid-March 2020, have you noticed any changes to your menstrual cycle? (yes/no/not applicable due to hormonal suppression/not applicable other)”, and participants that were postmenopausal were asked “Have you noticed any changes to your postmenopausal status since mid-March 2020? (yes/no).” Respondents who indicated changes to their menstrual cycle or postmenopausal status were prompted to check all that applied from a list of changes and provided with a free-text box (Survey available in Supplemental Section). All free-text responses were coded with thematic analysis using the methods outlined in Braun & Clarke (2006) to examine the potential variables influencing these changes^28^. All responses were assigned to one of two main themes: ‘Non-pandemic related’ and ‘Pandemic control measures.’ “Non-pandemic related” comments were obvious changes to their health and reproductive system at the start of the pandemic or preceding the pandemic. Alternatively, specific mentions of changes to their status or cycle, without medical reason, and stated as due to the pandemic were categorized as pandemic-related. After separating all responses into these two themes, sub-themes were created to identify reproductive changes that occurred during the pandemic. Sub-themes were created if one or more respondents expressed the same theme in their free-text response. Validated scores of general pandemic stress, anxiety, and depression were measured^29,30^. At the time of survey completion, participants were asked to recollect their mental health status during several pandemic phases that were based on the public health measures given at the time^1^ (Phase 1 lasted from mid-March 2020 to mid-May 2020, Phase 2 lasted from mid-May 2020 to mid-June 2020, Phase 3 lasted from mid-June 2020 until the end of November 2020. Phase 4 lasted from November 2020 to the date our survey closed). General pandemic stress was measured using the CoRonavIruS Health Impact Survey (CRISIS) V0.3. Participants were asked to self-report feelings of stress on a Likert scale from one (not at all) to five (extremely). Scores for this questionnaire range from 10–50 with higher scores indicating greater COVID-related stress.

Anxiety scores were calculated using the Generalized Anxiety Disorder questionnaire (GAD-7) The GAD-7 used self-reported feelings of anxiety on a Likert scale from zero (not at all) to three (nearly everyday) with scores ranging from 0–21. Scores above 10 suggest clinically significant levels of anxiety^31^. The Patient Health Questionnaire (PHQ-9) was used to measure self-reported symptoms of depression on a Likert scale from zero (not at all) to three (nearly everyday). Scores range from 0–27 with a score of 0–4 indicating minimal depression, 5–14 indicating mild to moderate depression and 15–27 indicating moderately severe to severe depression^1^. Internal consistency across data collection and Cronbach’s alpha in the current sample was very good (CRISIS: α = 0.882; GAD-7: 0.889, PHQ: α = 0.848).

### Statistical Analyses

Analyses were carried out using R v.4.1.3.^32^. The data set was divided into two separate groups: people with menstrual periods and those in menopause (postmenopause) as described above and summarized the percentages of those with any changes noted. We examined the relationship between mental health and changes in menopause status or menstruation by taking the average mental health scores across phases for each participant. These were entered in logistic regressions with change in menstruation or menopausal status as the outcome, and controlling for age, and presence of any coexisting chronic conditions (any of asthma, COPD, chronic lung disease, insulin resistance, diabetes, hypertension, heart disease, coronary artery disease, heart failure, cardiac arrhythmia, stroke, DVT, peripheral vascular disease, liver disease/cirrhosis, kidney disease, autoimmune disorder, pneumonia, or chronic neurologic or neuromuscular disorder). Significance was assessed using likelihood-ratio tests. Missing data were excluded from analyses.

## Results

### Survey Participants

Demographic information (age, ethnicity, gender, number of adults or children in the household, education, etc.) is seen in Table 1. Of the premenopausal participants, 98.4% identified as women, 0.1% identified as men and 1.5% as gender diverse. Of the postmenopausal participants, 99% identified as women, 0.1% identified as men and 0.8% as gender diverse. Given that nearly all females also identified as women, we have chosen to use the term women+ to refer to participants in our survey.

**Table 1.** Demographic characteristics of respondents. + denotes GenderQueer, Agender, Two-Spirit, other

| Menstruation Changes |  |  |  | Postmenopausal Changes |  |  |  |  |
| --- | --- | --- | --- | --- | --- | --- | --- | --- |
| Age | Total | No | Yes | P-value | Total | No | Yes | P-value |
|  | No. 1,866 | No. 1,347 | No. 519 |  | No. 2,305 | No. 2,150 | No. 155 |  |
| 25-29 | 183 (9.8%) | 127 (9.4%) | 56 (10.8%) | 0.14 | 5 (0.2%) | 4 (0.2%) | 1 (0.6%) | < 0.0001 |
| 30-39 | 627 (33.6%) | 446 (33.1%) | 181 (34.9%) |  | 38 (1.6%) | 34 (1.6%) | 4 (2.6%) |  |
| 40-49 | 808 (43.3%) | 602 (44.7%) | 206 (39.7%) |  | 174 (7.5%) | 151 (7.0%) | 23 (14.8%) |  |
| 50-59 | 242 (13.0%) | 166 (12.3%) | 76 (14.6%) |  | 948 (41.1%) | 854 (39.7%) | 94 (60.6%) |  |
| 60-69 | 6 (0.3%) | 6 (0.4%) | 0 (0.0%) |  | 1,140 (49.5%) | 1,107 (51.5%) | 33 (21.3%) |  |
| Gender |  |  |  |  |  |  |  |  |
| Woman | 1,837 (98.4%) | 1,333(99.0%) | 504 (97.1%) | 0.006 | 2,283 (99.0%) | 2,130 (99.1%) | 153 (98.7%) | 0.49 |
| Man | 1 (0.1%) | 1 (0.1%) | 0 (0.0%) |  | 3 (0.1%) | 3 (0.1%) | 0 (0.0%) |  |
| Non-Binary, GenderQueer, Agender, Two-spirit or other | 28 (1.5%) | 13 (1.0%) | 15 (2.9%) |  | 19 (0.8%) | 17 (0.8%) | 2 (1.3%) |  |
| Indigenous |  |  |  |  |  |  |  |  |
| Indigenous | 72 (3.9%) | 47 (3.5%) | 25 (4.8%) | 0.35 | 52 (2.3%) | 47 (2.2%) | 5 (3.2%) | 0.047 |
| Not Indigenous | 1,706 (91.4%) | 1,235 (91.7%) | 471 (90.8%) |  | 2,147 (93.1%) | 2,008 (93.4%) | 139 (89.7%) |  |
| Prefer not to answer | 19 (1.0%) | 15 (1.1%) | 4 (0.8%) |  | 13 (0.6%) | 10 (0.5%) | 3 (1.9%) |  |
| Missing | 69 (3.7%) | 50 (3.7%) | 19 (3.7%) |  | 93 (4.0%) | 85 (4.0%) | 8 (5.2%) |  |
| Latin American |  |  |  |  |  |  |  |  |
| Latin American | 40 (2.1%) | 25 (1.9%) | 15 (2.9%) | 0.21 | 21 (0.9%) | 18 (0.8%) | 3 (1.9%) | 0.16 |
| Not | 1,819 (97.5%) | 1,316 (97.7%) | 503 (96.9%) |  | 2,267 (98.4%) | 2,118 (98.5%) | 149 (96.1%) |  |
| Missing | 7 (0.4%) | 6 (0.4%) | 1 (0.2%) |  |  | 14 (0.7%) | 3 (1.9%) |  |
|  |  |  |  |  | 17 (0.7%) |  |  |  |
| **South Asian** |  |  |  |  |  |  |  |  |
| Not | 1,800 (96.5%) | 1,295 (96.1%) | 505 (97.3%) | 0.38 | 2,258 (98.0%) | 2,112 (98.2%) | 146 (94.2%) | 0.012 |
| South Asian | 59 (3.2%) | 46 (3.4%) | 13 (2.5%) |  | 30 (1.3%) | 24 (1.1%) | 6 (3.9%) |  |
| Missing | 7 (0.4%) | 6 (0.4%) | 1 (0.2%) |  | 17 (0.7%) | 14 (0.7%) | 3 (1.9%) |  |
| **Black** |  |  |  |  |  |  |  |  |
| Black | 19 (1.0%) | 15 (1.1%) | 4 (0.8%) | 0.61 | 8 (0.3%) | 8 (0.4%) | 0 (0.0%) | 1 |
| Not Black | 1,840 (98.6%) | 1,326 (98.4%) | 514 (99.0%) |  | 2,280 (98.9%) | 2,128 (99.0%) | 152 (98.1%) |  |
| Missing | 7 (0.4%) | 6 (0.4%) | 1 (0.2%) |  | 17 (0.7%) | 14 (0.7%) | 3 (1.9%) |  |
| **White** |  |  |  |  |  |  |  |  |
| Non-White | 368 (19.7%) | 281 (20.9%) | 87 (16.8%) | 0.044 | 245 (10.6%) | 222 (10.3%) | 23 (14.8%) | 0.077 |
| White | 1,491 (79.9%) | 1,060 (78.7%) | 431 (83.0%) |  | 2,043 (88.6%) | 1,914 (89.0%) | 129 (83.2%) |  |
| Missing | 7 (0.4%) | 6 (0.4%) | 1 (0.2%) |  | 17 (0.7%) | 14 (0.7%) | 3 (1.9%) |  |
| **Education** |  |  |  |  |  |  |  |  |
| More than high school | 1,705 (91.4%) | 1,232 (91.5%) | 473 (91.1%) | 0.93 | 1,924 (83.5%) | 1,788 (83.2%) | 136 (87.7%) | 0.18 |
| High school or less | 159 (8.5%) | 114 (8.5%) | 45 (8.7%) |  | 377 (16.4%) | 358 (16.7%) | 19 (12.3%) |  |
| Missing | 2 (0.1%) | 1 (0.1%) | 1 (0.2%) |  | 4 (0.2%) | 4 (0.2%) | 0 (0.0%) |  |
| **Number of Adults in Household** |  |  |  |  |  |  |  |  |
| One | 388 (20.8%) | 273 (20.3%) | 115 (22.2%) | 0.65 | 649 (28.2%) | 601 (28.0%) | 48 (31.0%) | 0.51 |
| Three or more | 364 (19.5%) | 263 (19.5%) | 101 (19.5%) |  | 526 (22.8%) | 488 (22.7%) | 38 (24.5%) |  |
| Two | 1,113 (59.6%) | 810 (60.1%) | 303 (58.4%) |  | 1,121 (48.6%) | 1,052 (48.9%) | 69 (44.5%) |  |
| Missing | 1 (0.1%) | 1 (0.1%) | 0 (0.0%) |  | 9 (0.4%) | 9 (0.4%) | 0 (0.0%) |  |
| **Children < 5** |  |  |  |  |  |  |  |  |
| None | 1,514 (81.1%) | 1,079 (80.1%) | 435 (83.8%) | 0.095 | 2,225 (96.5%) | 2,076 (96.6%) | 149 (96.1%) | 0.39 |
| One | 239 (12.8%) | 185 (13.7%) | 54 (10.4%) |  | 23 (1.0%) | 22 (1.0%) | 1 (0.6%) |  |
| Two or more |  |  |  |  |  |  |  |  |
|  | 70 (3.8%) | 54 (4.0%) | 16 (3.1%) |  | 14 (0.6%) | 12 (0.6%) | 2 (1.3%) |  |
| Missing | 43 (2.3%) | 29 (2.2%) | 14 (2.7%) |  | 43 (1.9%) | 40 (1.9%) | 3 (1.9%) |  |
| **Children 5-17** |  |  |  |  |  |  |  |  |
| None | 996 (53.4%) | 698 (51.8%) | 298 (57.4%) | 0.02 | 1,935 (83.9%) | 1,819 (84.6%) | 116 (74.8%) | 0.004 |
| One | 379 (20.3%) | 271 (20.1%) | 108 (20.8%) |  | 201 (8.7%) | 182 (8.5%) | 19 (12.3%) |  |
| Two or more | 464 (24.9%) | 357 (26.5%) | 107 (20.6%) |  | 121 (5.2%) | 105 (4.9%) | 16 (10.3%) |  |
| Missing | 27 (1.4%) | 21 (1.6%) | 6 (1.2%) |  | 48 (2.1%) | 44 (2.0%) | 4 (2.6%) |  |
| **Chronic Health Conditions** |  |  |  |  |  |  |  |  |
| None | 1,026 (55.0%) | 776 (57.6%) | 250 (48.2%) | 0.0003 | 998 (43.3%) | 944 (43.9%) | 54 (34.8%) | 0.029 |
| One or more | 839 (45.0%) | 570 (42.3%) | 269 (51.8%) |  | 1,300 (56.4%) | 1,199 (55.8%) | 101 (65.2%) |  |
| Missing | 1 (0.1%) | 1 (0.1%) | 0 (0.0%) |  | 7 (0.3%) | 7 (0.3%) | 0 (0.0%) |  |
| **Crisis score phase 1** |  |  |  |  |  |  |  |  |
| Mean (SD) | 27.8 ($\pm 8.0$) | 26.6 ($\pm 7.7$) | 30.9 ($\pm 8.0$) | < 0.0001 | 24.9 ($\pm 7.6$) | 24.6 ($\pm 7.4$) | 28.9 ($\pm 8.8$) | < 0.0001 |
| Missing | 63 (3.4%) | 51 (3.8%) | 12 (2.3%) |  | 86 (3.7%) | 78 (3.6%) | 8 (5.2%) |  |
| **Anxiety GAD-7 phase 1** |  |  |  |  |  |  |  |  |
| Mean (SD) | 6.9 ($\pm 5.2$) | 6.2 ($\pm 4.9$) | 8.7 ($\pm 5.5$) | < 0.0001 | 5.0 ($\pm 4.7$) | 4.8 ($\pm 4.6$) | 7.4 ($\pm 5.5$) | < 0.0001 |
| Missing | 49 (2.6%) | 40 (3.0%) | 9 (1.7%) |  | 106 (4.6%) | 100 (4.7%) | 6 (3.9%) |  |
| **Depression PHQ-9 phase 1** |  |  |  |  |  |  |  |  |
| Mean (SD) | 6.5 ($\pm 5.2$) | 5.7 ($\pm 4.7$) | 8.6 ($\pm 5.9$) | < 0.0001 | 5.0 ($\pm 4.5$) | 4.8 ($\pm 4.4$) | 7.8 ($\pm 5.7$) | < 0.0001 |
| Missing | 61 (3.3%) | 51 (3.8%) | 10 (1.9%) |  | 144 (6.2%) | 134 (6.2%) | 10 (6.5%) |  |

### Changes in menstrual symptoms

Among our sample of premenopausal women+, 519 (27.8% of the sample without suppressed cycles) reported that they had noticed changes in their menstrual cycle since March 2020. Of these 519 respondents, 44.3% indicated that their periods were more symptomatic (painful, more bleeding, etc.), 25.4%% indicated that their periods were longer, 23.7%% said they were having fewer periods than before the pandemic, 21.8% noticed having more periods than normal, 15.7% said their periods had gotten shorter, 2.3% said their periods were less symptomatic. Options were not mutually exclusive, and we received 784 answers from a potential 519 respondents. Moreover, 17.8% used free-text space to explain the changes to their menstrual cycles that they had noticed. The analysis of the free text responses indicated that there were twice as many respondents that indicated their changes in symptoms were due to the *Pandemic control measures*(Table 2). As those aged >40 years are more likely to be experiencing perimenopause, which could create menstrual disturbances independent of psychosocial stress, we undertook a sensitivity analysis of those who were less than 40 years old. Of this subset (n=810), 29.2% (n=237) reported changes in their menstrual cycle since the pandemic began. In addition, 52.7% indicated that their periods were more symptomatic (painful, more bleeding, etc.), 32.1% indicated that their periods had gotten longer, 13.1% said they were having fewer periods than normal, 20.7% noticed having more periods than normal, 19.0% said their periods had gotten shorter, 2.1% said their periods were less symptomatic, and 19.8% used free-text space to explain the changes to their menstrual cycles that they had noticed. Options were not mutually exclusive and we received 378 answers from a potential 237 respondents.

**Table 2.**
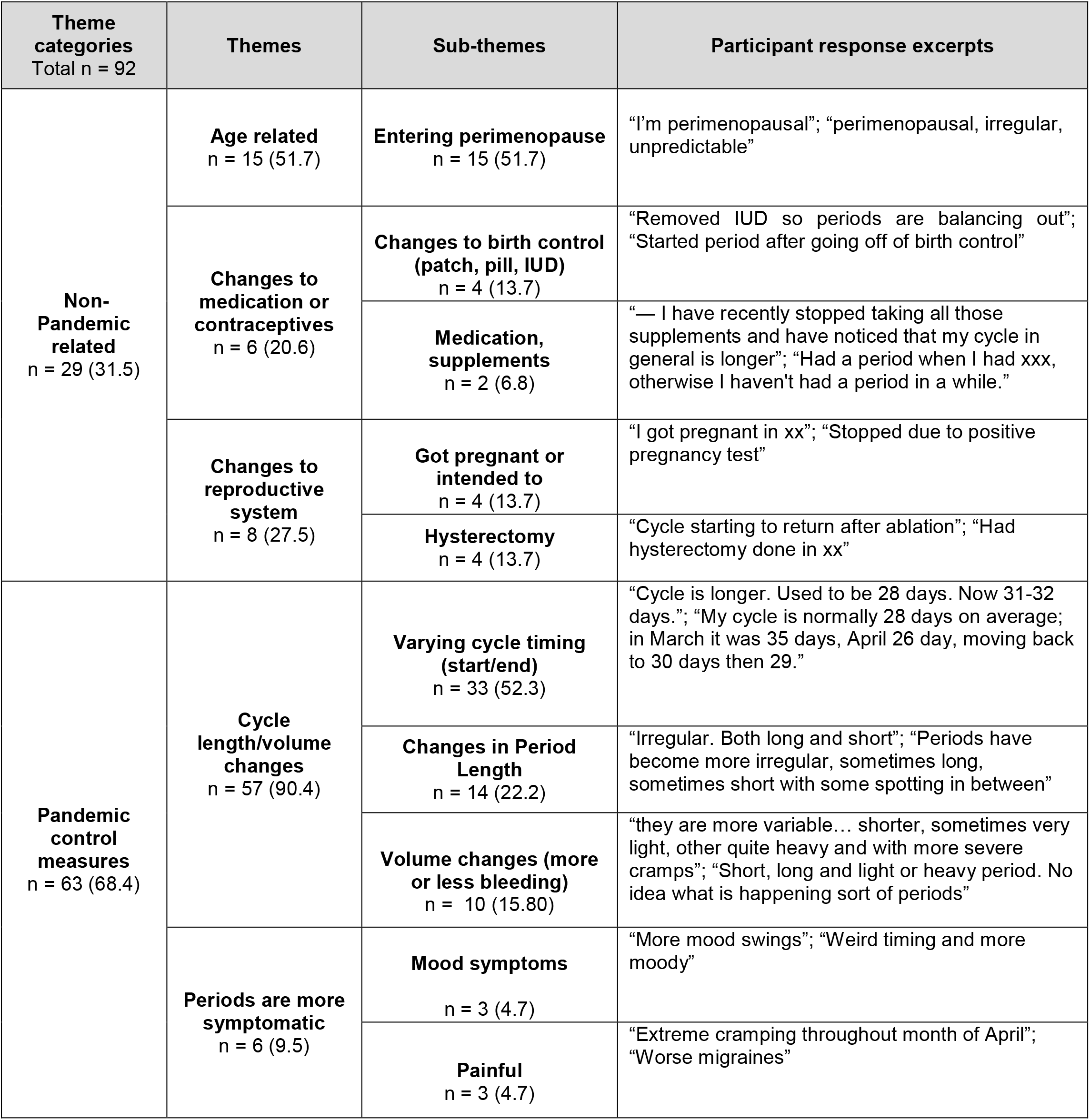
Summary of free-text responses for changes noticed in menstrual cycles. Sample sizes are given and percentages of total sample are given in parentheses.

### Changes in menopausal symptoms

Among our sample of postmenopausal females (n = 2305), 155 (6.7%) reported that they noticed changes in their postmenopausal status since mid-March 2020. Of this subset (n=155), 16.1% indicated that they started bleeding again and 12.3% indicated that they were experiencing “more menstrual symptoms”. Additionally, 72.9% used free-text space to explain the changes to their postmenopausal status. The analysis of the free-text responses revealed that there were four times as many respondents that indicated changes to postmenopausal status falling into the “due to the pandemic control measures” category (Table 3). As those aged <50 years are more likely to be experiencing perimenopause, which could create menopausal disturbances independent of psychosocial stress, a sensitivity analysis that restricted to those ≥50 (n = 1978) was conducted. Of this subset, there were 127 (6.4%) who reported that they had noticed changes in their postmenopausal status since before the pandemic. Of this subset (n=127), 18.1% indicated that they started bleeding again and 7.9% indicated that they were experiencing “more menstrual symptoms”. Additionally, 71.7% of this subset used free-text space to explain the changes to their postmenopausal status that they had noticed. Analysis of these free-text responses revealed that 19.8% of reported changes were associated with the onset of the pandemic and 80.1% were not not pandemic-related.

**Table 3.**
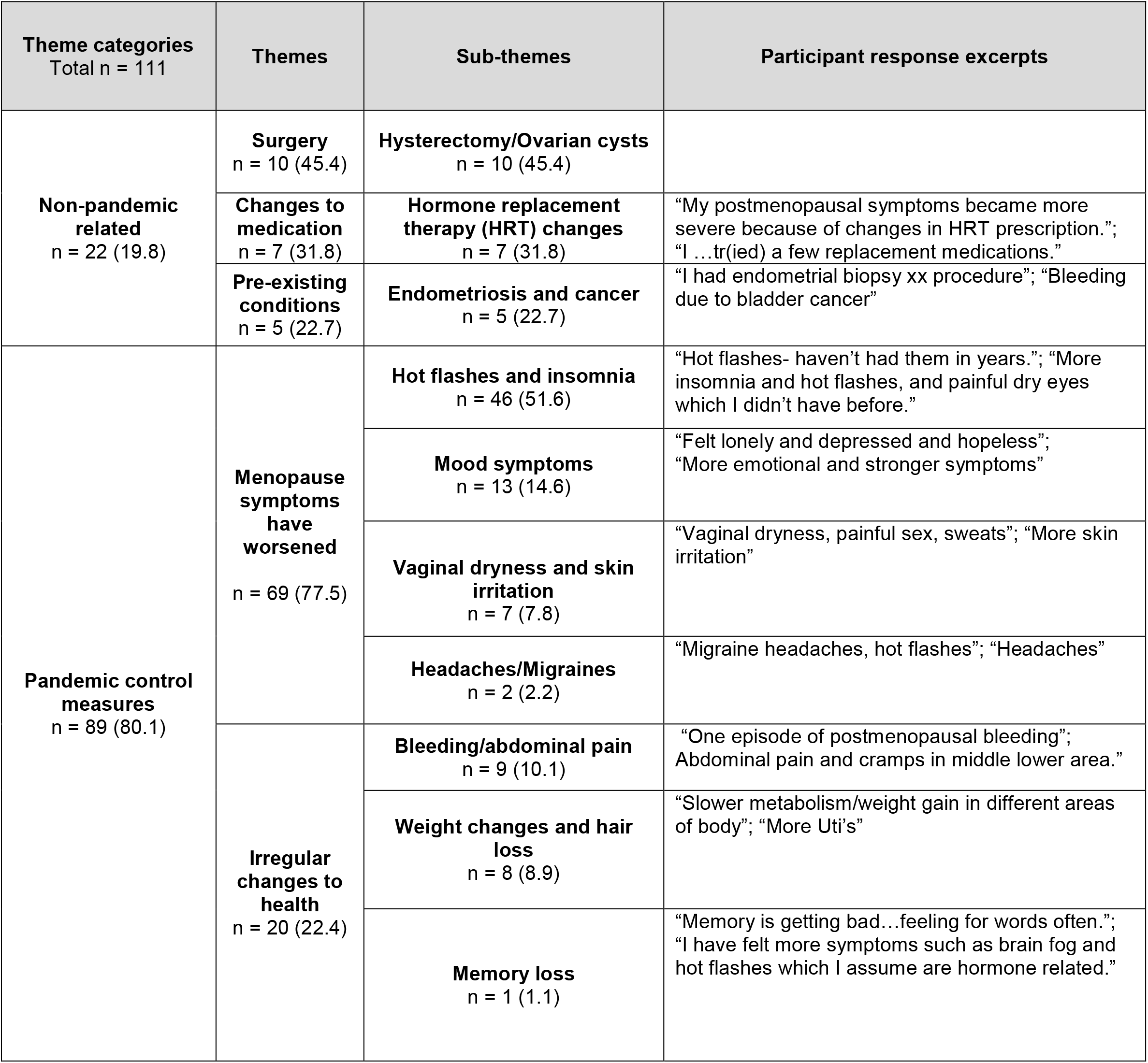
Summary of free-text responses for changes noticed in postmenopausal status. Sample sizes are given and percentages of total sample are given in parentheses.

| Theme categories<br>Total n = 111 | Themes | Sub-themes | Participant response excerpts |
| --- | --- | --- | --- |
| **Non-pandemic related**<br>n = 22 (19.8) | **Surgery**<br>n = 10 (45.4) | **Hysterectomy/Ovarian cysts**<br>n = 10 (45.4) |  |
|  | **Changes to medication**<br>n = 7 (31.8) | **Hormone replacement therapy (HRT) changes**<br>n = 7 (31.8) | "My postmenopausal symptoms became more severe because of changes in HRT prescription."; "I ...tr(ied) a few replacement medications." |
|  | **Pre-existing conditions**<br>n = 5 (22.7) | **Endometriosis and cancer**<br>n = 5 (22.7) | "I had endometrial biopsy xx procedure"; "Bleeding due to bladder cancer" |
| **Pandemic control measures**<br>n = 89 (80.1) | **Menopause symptoms have worsened**<br>n = 69 (77.5) | **Hot flashes and insomnia**<br>n = 46 (51.6) | "Hot flashes- haven't had them in years."; "More insomnia and hot flashes, and painful dry eyes which I didn't have before." |
|  |  | **Mood symptoms**<br>n = 13 (14.6) | "Felt lonely and depressed and hopeless"; "More emotional and stronger symptoms" |
|  |  | **Vaginal dryness and skin irritation**<br>n = 7 (7.8) | "Vaginal dryness, painful sex, sweats"; "More skin irritation" |
|  |  | **Headaches/Migraines**<br>n = 2 (2.2) | "Migraine headaches, hot flashes"; "Headaches" |
|  | **Irregular changes to health**<br>n = 20 (22.4) | **Bleeding/abdominal pain**<br>n = 9 (10.1) | "One episode of postmenopausal bleeding"; "Abdominal pain and cramps in middle lower area." |
|  |  | **Weight changes and hair loss**<br>n = 8 (8.9) | "Slower metabolism/weight gain in different areas of body"; "More Uti's" |
|  |  | **Memory loss**<br>n = 1 (1.1) | "Memory is getting bad...feeling for words often."; "I have felt more symptoms such as brain fog and hot flashes which I assume are hormone related." |

### Higher pandemic stress, anxiety, and depression symptoms were related to more disturbances in menstrual and menopausal symptoms

Across both groups, higher average scores on the psychosocial measures were associated with increased odds of disturbances in their cycles or changes to postmenopausal status. For premenopausal women+ the odds ratio (OR) for pandemic crisis score was 1.06 (95%CI: 1.04-1.08), suggesting that the odds of disturbance increased by 6% for every increase in 1 point along the crisis scale. Controlling for age and chronic conditions, the estimated marginal proportion with disturbances when crisis score = 10 was 30.5% (95%CI: 24.0%-38.0%), and when crisis score = 40 was 70.3% (95%CI:65.0%-75.2%) (Figure 1A). The OR for anxiety was 1.11 (95%CI: 1.08-1.14), suggesting that the odds of disturbance increased by 11% for every increase in 1 point along the GAD-7 scale. Controlling for age and chronic conditions, the estimated marginal proportion with disturbances when the GAD-7 score = 0 was 15.7% (95%CI: 13.2%-18.4%), and when the GAD-7 score = 10 was 34.8% (95%CI:32.0%-37.6%) (Figure 1B). The OR for depression was 1.13 (95%CI: 1.10-1.15), suggesting that the odds of disturbance increased by 13% for every increase in 1 point along the PHQ-9 scale. Controlling for age and chronic conditions, the estimated marginal proportion with disturbances when the PHQ-9 score = 0 was 14.5% (95%CI: 12.3%-17.1%), and when the PHQ-9 score = 12 was 41.9% (95%CI:38.2%-45.7%) (Figure 1C).

**Figure 1.**
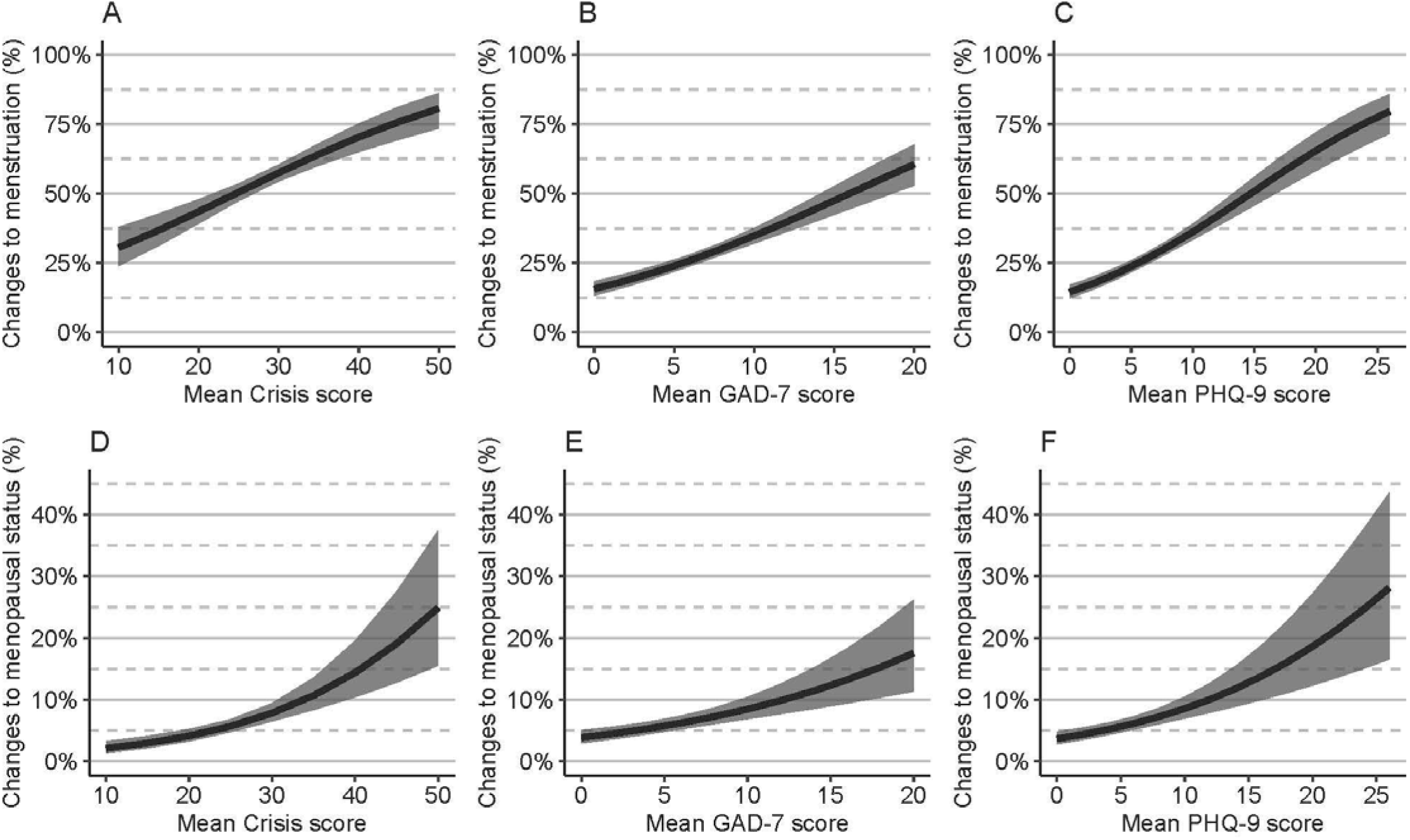
Changes to menstrual and menopausal status during early COVID-19 pandemic increased mean CoRonavIruS Health Impact survey (CRISIS) scores, anxiety (GAD-7) and depression symptoms (PHQ-9) in women+. Predicted marginal proportions and 95%CI from logistic regressions controlling for age and any comorbid chronic conditions. A) CRISIS scores in women+ by changes to menstrual status B) GAD-7 scores in women+ by changes to menstrual status C) PHQ-9 scores in women+ by changes to menstrual status D) CRISIS scores in women+ by changes to menopausal status E) GAD-7 scores in women+ by changes to menopausal status F) PHQ-9 scores in women+ by changes to menopausal status. CI=confidence interval.

For postmenopausal women+, the odds ratio (OR) for pandemic crisis score was 1.07 (95%CI: 1.04-1.10), suggesting that the odds of changes increased by 7% for every increase in 1 point along the crisis scale. Controlling for age and chronic conditions, the estimated marginal proportion with changes when crisis score = 10 was 2.1% (95%CI: 1.4%-3.3%), and when crisis score = 40 was 14.4% (95%CI:10.4%-19.6%) (Figure 1D). The OR for anxiety was 1.09 (95%CI: 1.05-1.12), suggesting that the odds of changes increased by 9% for every increase in 1 point along the GAD-7 scale. Controlling for age and chronic conditions, the estimated marginal proportion with changes when the GAD-7 score = 0 was 3.9% (95%CI: 3.0%-5.1%), and when the GAD-7 score = 10 was 8.5% (95%CI:6.9%-10.4%) (Figure 1E). The OR for depression was 1.09 (95%CI: 1.06-1.13), suggesting that the odds of changes increased by 9% for every increase in 1 point along the PHQ-9 scale. Controlling for age and chronic conditions, the estimated marginal proportion with changes when the PHQ-9 score = 0 was 3.7% (95%CI: 2.8%-4.8%), and when the PHQ-9 score = 12 was 10.1% (95%CI:7.9%-12.7%) (Figure 1F).

### Relationship of menstrual or postmenopausal disturbances to number of children

As previous studies have found an association between the number of children and stress^33^, we also examined whether the number of children influenced our results. We ran these analyses in premenstrual women+ that were under 40, choosing to exclude individuals who may have been perimenopausal based on age. There was no significant relationship between the number of children and whether the participant indicated they had disturbances to their menstrual cycle (p=0.100). We next examined whether there was a relationship between menopausal disturbances and the number of children. There was a significant relationship (p=0.007), however when we used age as a covariate the effect was no longer significant (p=0.409). There were also no significant relationships between the number of children and pandemic stress scores, after controlling for age and changes to menstruation or menopause (p=0.167 and 0.400, respectively).

## Discussion

In a sample of 4171 surveyed women+ in BC that met our inclusion criteria, 27.8% of naturally cycling women+ had a disruption in their menstrual cycle and 6.7% of postmenopausal women+ indicated a change in their status, across the first year of the COVID-19 pandemic and prior to widespread rollout of COVID-19 vaccines. These disturbances to the reproductive cycles were related to higher levels of anxiety, depression, and perceived stress, but not to the number of children, in both pre and postmenopausal women+. Women+ with higher stress scores were more likely to have experienced menstrual cycle phase disturbance with an approximate doubling in the estimated proportion between crisis scores of 10 (31%) compared to 35 (64%) for premenopausal women+, and a doubling of postmenopausal changes between crisis scores of 10 (2%) and 20 (4%) for postmenopausal women+. Similarly, higher levels of anxiety and depression were associated with higher proportions of menstrual phase disturbance and postmenopausal changes (Figure 1 A-F). We previously found that women had higher levels of perceived stress, depression, and anxiety compared to men, using data from the same source^1^. Our findings here indicate that both pre and postmenopausal women+ with higher levels of mental distress experienced greater disturbances in reproductive cycles. Stress has pervasive effects on mental and physical health and our results add to the growing data that female-specific reproductive cycles are also affected.

### During the first year of the pandemic menstrual cycle disturbances were related to distress, anxiety and depression

We found that in the first year of the pandemic, 27.8% of the naturally-cycling women+ surveyed had disturbances to their menstrual cycles. This is slightly lower than other surveys suggesting that 44% of women (including women that were younger than our own cohort 18-24) noticed changes in their cycles using fertility tracking devices^34,35^. Other studies have suggested that the baseline rates of menstrual disturbances are ∼15%^36,37^. Premenopausal women in our study with reported menstrual disturbances had 3.4 times higher perceived stress. Our findings are consistent with another study that showed an association between perceived stress during the COVID-19 pandemic and menstrual irregularities. Although the literature suggests that parental responsibility increases perceived stress in women compared to men^33^, we found no evidence that there was a significant relationship between the number of children, and changes in the menstrual cycle or in perceived stress in women+ younger than 40. Our data also indicate that higher anxiety scores and higher depression scores were associated with reported menstrual cycle disturbances.

### Increased menopause symptoms were associated with increased perceived stress, anxiety and depression

We also found that fewer than ten percent of postmenopausal women+ noticed changes to their menopausal status during the first year of the pandemic. Those who noticed changes in their status reported more menopausal symptoms, and furthermore these individuals were more likely to score higher on perceived stress, depression and anxiety. Perimenopause is thought to be a time of heightened risk for mental health disruption^38^ and those with menopausal symptoms may be at greater risk to develop psychiatric disorders^39^. The percentage of disturbances in reproductive symptoms was four times greater in our sample of premenopausal (27.8%) compared to postmenopausal women+ (6.7%). It is not clear why this large difference occurred but may be due to age, influence of stress on the HPG axis, or level of stress overall. Indeed, other studies, including data from this same sample, have found that perceived stress, anxiety and depression scores were reduced with increasing age^1,40^. Thus, our findings of greater premenopausal versus postmenopausal disturbances aligns with data indicating mental health indices were lower for older adults during the first year of the pandemic^1^. Hypothalamic-pituitary-adrenal (HPA) axis activation, via stress, initially stimulates, and chronic activation inhibits the HPG resulting in menstrual abnormalities^41^ and as these interactions vary across age^42,43^, this may also explain our findings as there would have been less HPA activation in the postmenopausal women+ in our sample. However, it is also possible that the premenopausal versus postmenopausal difference was due, in part, to the differences in the question posed to the two different groups, as our survey asked about menopause status rather than symptoms of menopause.

### Relevance and Limitations

Allostatic load, or the cumulative burden of a variety of stressors, affects cardiovascular and metabolic health^44,45^. However, it is important to acknowledge that female-specific factors, such as menstruation and menopause, are also impacted by chronic stress. Others have postulated that female health is cyclical as menstrual patterns correspond to changes in immune system function^12^. Moreover, there are specific interactions between gonadal hormones and immune cells^46^ emphasizing important crosstalk^47^. It is possible that the mechanism behind stress and menstrual changes also involves glucocorticoid receptors in the endometrium^48^. Although SARS-CoV-2 infection and vaccines use can also disrupt reproductive cycles in the short term^7,10^, we do not believe this influenced our findings because we found that the seroprevalence of previous infection in this cohort was 2.9%^49^ and vaccines were not widely available to the general BC population at the time of this survey. There are limitations to our study. This was a retrospective survey that required online access. However, our findings of high levels of menstrual disturbances is consistent with other studies using data from menstrual tracking apps^34^. In our survey we did not obtain information on the normal levels of menstrual or menopausal symptoms pre-COVID19 pandemic hence we are only able to cross study comparisons.

This survey was limited to residents of the province of BC and our unique patterns of pandemic restriction measures may have impacted results. We also asked about menopausal status rather than menopausal symptoms and this may have reduced responses to this population of postmenopausal people. In our survey we also did not clarify why people had suppressed cycles (e.g which type of hormonal contraceptives, gender affirming hormone therapy) which future studies could explore. We cannot rule out a selection bias in this study, and it is possible that individuals who experienced higher stress pre- and post pandemic were more likely to participate. Similarly, those with menstrual abnormalities may be hyper-aware of their menstrual cycle and led them to participate in the current study.

## Conclusions

The stress associated with the pandemic has impacted both our physical and mental health and our findings suggest that this includes female-specific physical health characteristics. It is imperative to continue advancing the literature on external and internal factors impacting women’s reproductive health. Factors that can mitigate against the effects of stress such as social support^50^ and exercise^51^, which were limited during the initial phases of the pandemic, may have exacerbated the negative outcomes on reproductive cycles for women+. Given the lack of attention to women’s health data^52^, more studies examining female-specific health indices are needed in the literature.

## Data Availability

Data cannot be shared publicly because of ethical restrictions. Data are available from the UBC Research Ethics Board (contact via) for researchers who meet the criteria for access to confidential data.

## Acknowledgements

We thank David M. Goldfarb and Melanie Murray for their contributions to this work.

No prior publication or abstract presentations of this work.

## Supplement

1. Are you postmenopausal (have not had a period in 1 year or more)?
  - Yes
  - No
2. [If “No” to question 1] Since the COVID-19 pandemic and subsequent public health measures in mid-March 2020, have you noticed any changes to your menstrual cycle?
  - Yes
  - No
3. Not applicable: I have a suppressed menstrual cycle due to hormonal contraception, breast feeding, a medical condition, etc.
4. Not applicable: other
5. [If “yes” to question 2] What changes have you noticed to your menstrual cycle?
  - Periods have gotten shorter
  - Periods have gotten longer
  - Periods are more symptomatic (painful, more bleeding, etc.)
  - Periods are less symptomatic
  - More periods than normal
  - Fewer periods than normal
  - Other
6. [If “other” to question 3] What other changes in your menstrual cycle have you noticed since COVID-19?
7. [If “yes” to question 1] Have you noticed any changes to your postmenopausal status since mid-March 2020?
  - Yes
  - No
8. [If “yes” to question 5] What changes have you noticed?
  - Started bleeding again
  - More menstrual symptoms
  - Other
9. [If “other” to question 6] Please specify the “Other” changes you have experienced in your postmenopausal status

**Menstrual disturbances**

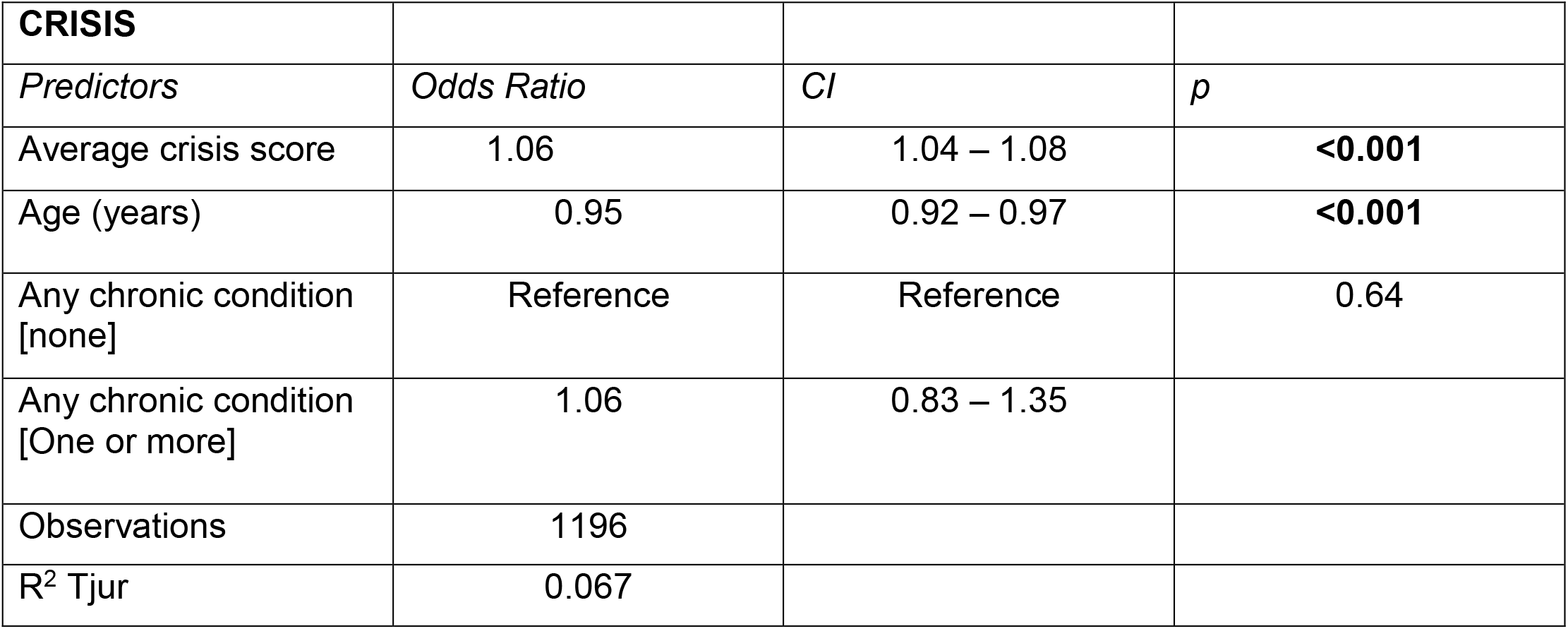

**Estimated marginal predictions from the model averaged over chronic conditions and age**

| **CRISIS Score** | **Predicted marginal proportion** | **95% CI** |
| --- | --- | --- |
| **10** | 0.31 | [0.24, 0.38] |
| **15** | 0.37 | [0.31, 0.43] |
| **20** | 0.43 | [0.39, 0.48] |
| **25** | 0.5 | [0.47, 0.54] |
| **30** | 0.57 | [0.54, 0.60] |
| **35** | 0.64 | [0.60, 0.68] |
| **40** | 0.7 | [0.65, 0.75] |
| **50** | 0.81 | [0.73, 0.86] |

| **GAD-7** |  |  |  |
| --- | --- | --- | --- |
| *Predictors* | *Odds Ratio* | *CI* | *p* |
| Average GAD-7 score | 1.11 | 1.09 – 1.14 | **<0.001** |
| Age (years) | 1.01 | 0.99 – 1.02 | 0.292 |
| Any chronic condition<br>[none] | Reference | Reference | 0.021 |
| Any chronic condition<br>[One or more] | 1.29 | 1.04 – 1.59 |  |
| Observations | 1805 |  |  |
| R<sup>2</sup> Tjur | 0.054 |  |  |

**Estimated marginal predictions from the model averaged over chronic conditions and age**

| **GAD-7 score** | **Predicted marginal proportion** | **95% CI** |
| --- | --- | --- |
| **0** | 0.16 | [0.13, 0.18] |
| **2** | 0.19 | [0.16, 0.21] |
| **6** | 0.26 | [0.24, 0.28] |
| **8** | 0.3 | [0.28, 0.32] |
| **10** | 0.35 | [0.32, 0.38] |
| **12** | 0.4 | [0.36, 0.43] |
| **14** | 0.45 | [0.40, 0.50] |
| **20** | 0.61 | [0.53, 0.68] |

| **PHQ-9** |  |  |  |
| --- | --- | --- | --- |
| *Predictors* | *Odds Ratio* | *CI* | *p* |
| Average PHQ-9 score | 1.13 | 1.10 – 1.15 | **<0.001** |
| Age (years) | 1.01 | 0.99 – 1.02 | 0.292 |
| Any chronic condition<br>[none] | Reference | Reference | 0.021 |
| Any chronic condition<br>[One or more] | 1.29 | 0.98 – 1.51 |  |
| Observations | 1794 |  |  |
| R<sup>2</sup> Tjur | 0.074 |  |  |

**Estimated marginal predictions from the model averaged over chronic conditions and age**

| **PHQ-9 score** | **Predicted marginal proportion** | **95% CI** |
| --- | --- | --- |
| **0** | 0.15 | [0.12, 0.17] |
| **4** | 0.22 | [0.19, 0.24] |
| **6** | 0.26 | [0.24, 0.28] |
| **10** | 0.36 | [0.33, 0.39] |
| **14** | 0.48 | [0.43, 0.53] |
| **16** | 0.54 | [0.48, 0.59] |
| **20** | 0.65 | [0.58, 0.72] |
| **26** | 0.8 | [0.72, 0.86] |

**Post-menopausal changes**

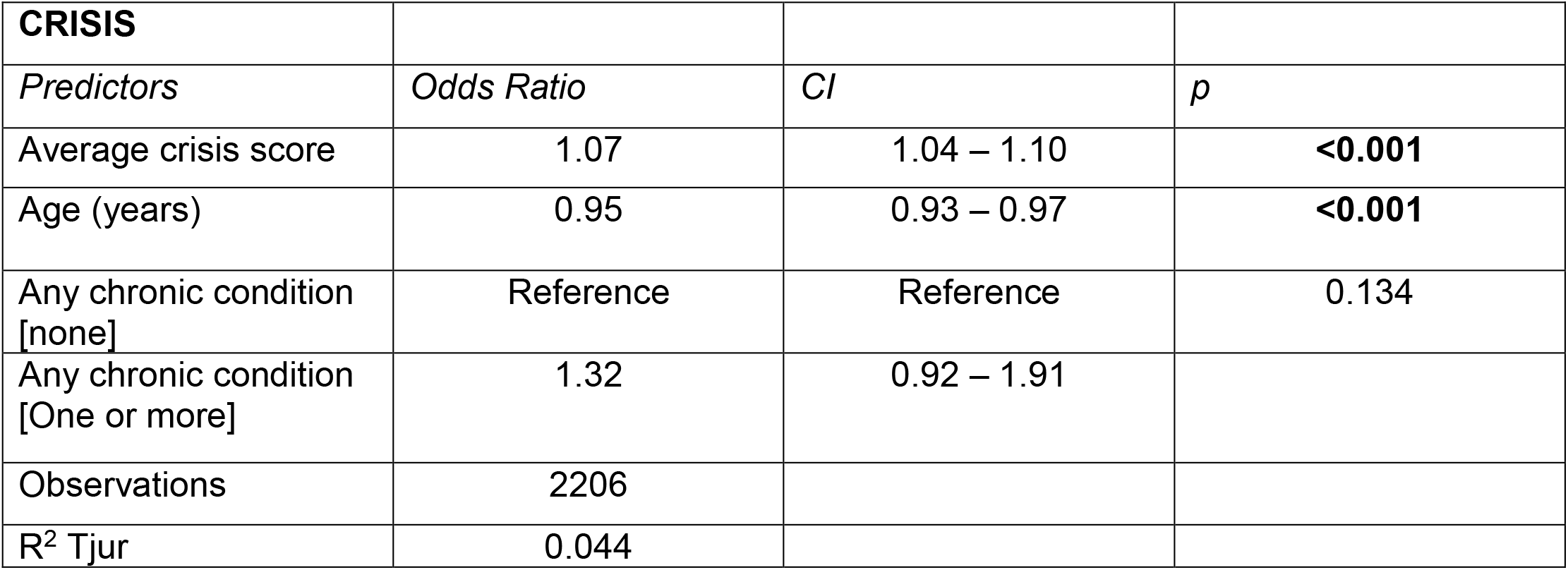

| **CRISIS** |  |  |  |
| --- | --- | --- | --- |
| *Predictors* | *Odds Ratio* | *CI* | *p* |
| Average crisis score | 1.07 | 1.04 – 1.10 | **<0.001** |
| Age (years) | 0.95 | 0.93 – 0.97 | **<0.001** |
| Any chronic condition<br>[none] | Reference | Reference | 0.134 |
| Any chronic condition<br>[One or more] | 1.32 | 0.92 – 1.91 |  |
| Observations | 2206 |  |  |
| R<sup>2</sup> Tjur | 0.044 |  |  |

**Estimated marginal predictions from the model averaged over chronic conditions and age**

| **CRISIS Score** | **Predicted marginal proportion** | **95% CI** |
| --- | --- | --- |
| **10** | 0.02 | [0.01, 0.03] |
| **15** | 0.03 | [0.02, 0.04] |
| **20** | 0.04 | [0.03, 0.05] |
| **25** | 0.06 | [0.05, 0.07] |
| **30** | 0.08 | [0.07, 0.09] |
| **35** | 0.11 | [0.08, 0.14] |
| **40** | 0.14 | [0.10, 0.20] |
| **50** | 0.25 | [0.16, 0.37] |

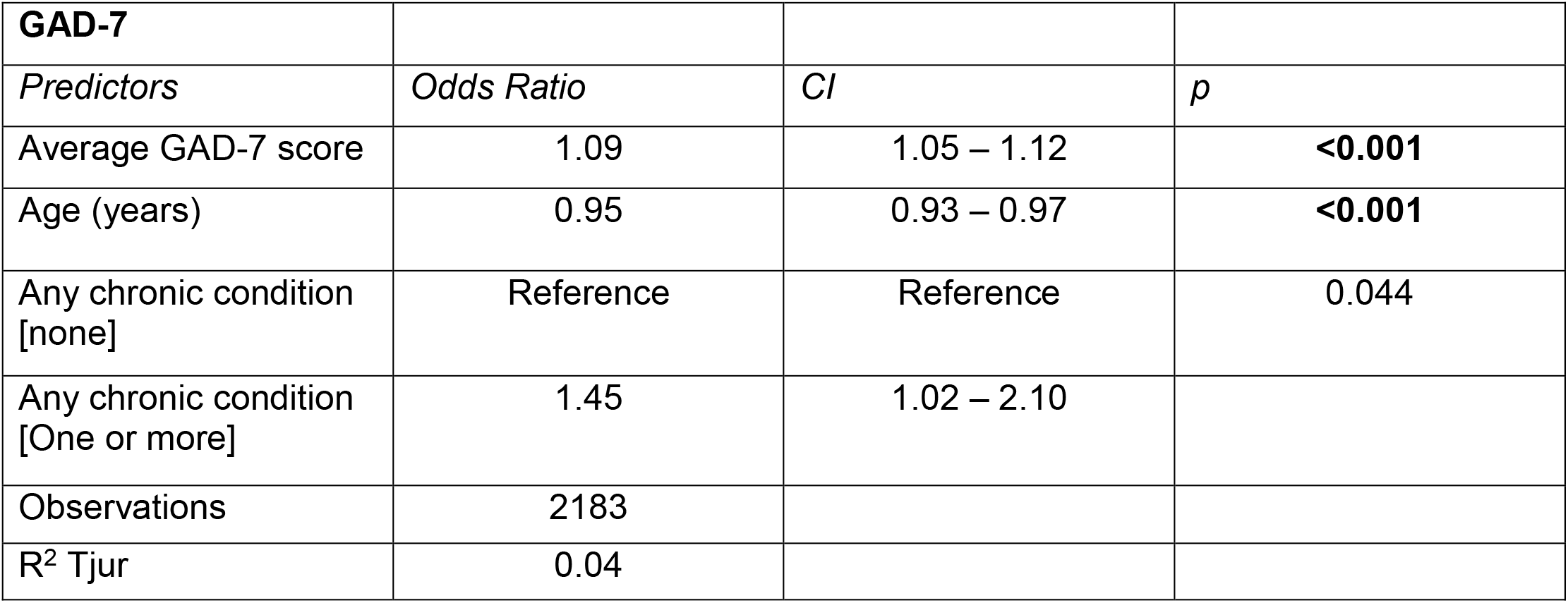

| **GAD-7** |  |  |  |
| --- | --- | --- | --- |
| *Predictors* | *Odds Ratio* | *CI* | *p* |
| Average GAD-7 score | 1.09 | 1.05 – 1.12 | **<0.001** |
| Age (years) | 0.95 | 0.93 – 0.97 | **<0.001** |
| Any chronic condition<br>[none] | Reference | Reference | 0.044 |
| Any chronic condition<br>[One or more] | 1.45 | 1.02 – 2.10 |  |
| Observations | 2183 |  |  |
| R<sup>2</sup> Tjur | 0.04 |  |  |

**Estimated marginal predictions from the model averaged over chronic conditions and age**

| **GAD-7 score** | **Predicted marginal proportion** | **95% CI** |
| --- | --- | --- |
| **0** | 0.04 | [0.03, 0.05] |
| **2** | 0.05 | [0.04, 0.06] |
| **6** | 0.06 | [0.05, 0.07] |
| **8** | 0.07 | [0.06, 0.09] |
| **10** | 0.08 | [0.07, 0.10] |
| **12** | 0.1 | [0.08, 0.13] |
| **14** | 0.11 | [0.09, 0.15] |
| **20** | 0.18 | [0.11, 0.26] |

| **PHQ-9** |  |  |  |
| --- | --- | --- | --- |
| *Predictors* | *Odds Ratio* | *CI* | *p* |
| Average PHQ-9 score | 1.09 | 1.06 – 1.13 | **<0.001** |
| Age (years) | 0.95 | 0.93 – 0.97 | **<0.001** |
| Any chronic condition<br>[none] | Reference | Reference | 0.144 |
| Any chronic condition<br>[One or more] | 1.31 | 0.92 – 1.90 |  |
| Observations | 2158 |  |  |
| R<sup>2</sup> Tjur | 0.043 |  |  |

**Estimated marginal predictions from the model averaged over chronic conditions and age**

| **PHQ-9 score** | **Predicted marginal proportion** | **95% CI** |
| --- | --- | --- |
| **0** | 0.04 | [0.03, 0.05] |
| **4** | 0.05 | [0.04, 0.06] |
| **6** | 0.06 | [0.05, 0.07] |
| **10** | 0.09 | [0.07, 0.10] |
| **14** | 0.12 | [0.09, 0.16] |
| **16** | 0.14 | [0.10, 0.19] |
| **20** | 0.19 | [0.12, 0.27] |
| **26** | 0.28 | [0.17, 0.44] |

